# Whole-Genomic Analysis of Antimicrobial Resistance in *Campylobacter jejuni* in Ethiopia, Kenya and Tanzania: A One Health Approach

**DOI:** 10.64898/2025.12.29.25343130

**Authors:** Jacksoni J. Maleva, Veronica E. Linkanti, Misambwa A. Yongolo, Yohane D. Sebogo, Emiliana F. Mallya, Edson F. Kimario, Jacob G. Msafiri, Catheline T. Kameka, Charles N. Sekelwa, Violet M. Temba, Erick A. Msafiri, Ashraph H. Mwalim, Eliud B. Felcian, Emmanuel Mwampale, Fatma Rashid, Beatus Lyimo

## Abstract

*Campylobacter jejuni* is a zoonotic bacterium causing foodborne gastroenteritis in humans worldwide, whose main symptom is diarrhea. The infection is severe mostly in children and in immunocompromised individuals. Currently, the bacterium has become increasingly resistant to antibiotics, especially those first-choice drugs used to treat campylobacteriosis posing a significant health threat towards the treatment outcomes. The burden of campylobacteriosis and Antimicrobial Resistance (AMR) remains significant with limited genomic surveillance. This study aimed to characterize the resistome (ARGs), virulence factors as well as population structure across *Homo sapiens*, Milk (from dairy cattle), goat, *Bos indicus*, *Ovis aries*, and *Gallus gallus* in three countries Ethiopia, Kenya and Tanzania through the use of One Health Whole Genome Sequencing (WGS) approach.

A total of 161 *C. jejuni* publicly available WGS were retrieved from NCBI database and analyzed by using established WGS bioinformatics pipelines from genome assembly and annotation, AMR gene identification via ResFinder -ABRIcate, virulence genes were detected via ABRicate/ VFDB. Visualization of gene distribution and population structure were done using heatmap, Venn diagrams, principal component analysis and minimum spanning tree for comparative analysis.

Out of 161 *C. jejuni* WGS, 130 (80.75%) sequences were positive to one or more than one ARGs. Among detected ARGs, the resistome was dominated by β-lactam (blaOXA 193, blaOXA-61, blaOXA-184 and blaOXA-489) genes. Two genes linked to tetracycline resistance (tet(O/32/O), and tet(O) were found in Ethiopia and Tanzania while resistance to aminoglycoside ant (6)-Ia was the least detected. The *Gallus gallus-Homo sapiens* transmission (zoonotic transmission) was portrayed by the overlap of ARGs (blaOXA-193 and tet(O) and PCA clustering. The conserved virulence gene profiles were shared by all isolated (cadF, jlpA, cdtA, cdtB, cdtC and flagellar genes).

The present study adds to the current knowledge on molecular epidemiology and AMR development in *C. jejuni* species in Eastern African countries and globally. The findings underscore the need for sustained region-specific genome surveillance under One Health framework to inform AMR stewardship and public health interventions.

## INTRODUCTION

Campylobacteriosis, an infectious disease caused by bacteria of genus *Campylobacter* dominated by *C. jejuni* and *C. coli*. Globally, *Campylobacter* is one of the four causative agents of diarrheal diseases as the result of food contamination with 550 million people falling ill yearly (including 220 million children under the age of 5 years (WHO, 2020). It is a major global health challenge due to its widespread nature, zoonotic nature as well as increasing resistance to various antibiotics (WHO, 2020). Africa is estimated to have the world’s highest incidence of campylobacteriosis from human and animals (French et al, 2024), though the burden of the disease remains poorly documented and under recognized due to poor national surveillance programs established compared to high income countries (Oberhelman A and Taylor N, 2000).

In low and middle income countries (LMICs) were sanitation is poor, limited water and food safety, the risks of human *Campylobacter* infections is high (Lee et al., 2013). Most outbreaks are caused by consumption of poultry meats from laying hens, turkeys, ostriches, ducks and broilers as well as poultry-derived products (Taylor et al., 2013; Epps et al., 2013). Poultry, especially broiler chicken are major reservoir of *campylobacte*r spp., particularly *C. jejuni*, because their intestinal system provides an ideal biological habitat for the survival and growth of these bacteria owing to their high body temperature (Ammar, 2021; EFSA, 2021). Gahamanyi et al., (2020) estimated that, the prevalence of *Campylobacter* in Sub Saharan Africa ranges from 1.7 to 62.7% in humans and 1.2–80% in animals. Also, ruminant animals such as pigs, sheep, goat and cows contribute to environmental contamination through fecal shedding.

De Vries et al, (2018) and Seguino et al., (2018) highlighted that farm animals are potential reservoirs of *Campylobacter* spp. which causing gastrointestinal infections in human (de Vries et al, 2018). They have also been reported to be involved in extra gastrointestinal manifestations, including bacteremia, lung infections, brain abscesses, meningitis, and reactive arthritis, in individual cases and small cohorts of patients (Man M 2011). In many developing countries such as Tanzania, smallholder livestock keeping closely to households and minimal biosecurity practices increases the direct transmission rate (Roth, 2023). This dynamic interface between human, animals and environment plays a key role in transmission of *Campylobacter* spp.

Antimicrobials including antibiotics, antifungals and antiparasitic are used for treatment and/ or prevention of infections (WHO,2021). When microorganisms do not respond to the antimicrobials that where once effective in controlling infection, they as said to be resistance to antimicrobials.

According to the World Health Organization (WHO), antimicrobial resistance (AMR) continues to pose a major global public health threat and has been included in global health priorities following recent evaluations (WHO, 2023). AMR undermines the effectiveness of life-saving treatments and makes previously treatable infections increasingly difficult, or in some cases impossible, to cure, thereby threatening the quality and safety of health care systems worldwide (WHO, 2023). Recent evidence from the Global Antimicrobial Resistance and Use Surveillance System (GLASS) indicates the resistance rise in many common bacterial pathogens, with substantial increases observed across several pathogen–antibiotic combinations (WHO, 2025). Furthermore, WHO warns that the continued spread of antibiotic resistance highlights the urgent need for coordinated global action to strengthen surveillance, improve antimicrobial stewardship, expand access to diagnostics, and accelerate the development of new antimicrobial agents (WHO, 2025). Globally AMR complications have associated with 4.7 million with bacterial AMR responsible for about 1.14 million deaths annually (Naghavi et al., 2024). The AMR commissioned by the United Kingdom government forecasted that, if there will be no joint action plan for antimicrobial resistance mitigation it could kill 10 million people per year by 2050 (Murray et al.,2022). The burden of the disease is worse in LMICs, attributed by poor surveillance systems, improper use of antibiotics in both human and animals, limited experts and diagnostic capacity (Laxminarayan et al.,2016). Children under five years have become more susceptible to AMR due to high morbid and mortality of about 99.65% in LMICs (Laxminarayan et al.,2016; Baker et al.,2018: UNICEF, 2023).

Moreover, AMR is also a growing problem in African livestock production systems, posing a threat to human and animal health and the associated economic value chain (Kauthar M et al 2024). In livestock sector, antimicrobials are mainly used for the treatment of animal infections and growth promotion, but uncontrolled, misuse and over use as well as sub therapeutic doses promote the rise of antimicrobial resistance (Ma et al., 2021). Antibiotics targeting bacteria, continue to be unresponsive in treatment of bacterial infection due to the existence of AMR genes, with cattle and poultry serving as important reservoir for these genes (Zalewska et al 2021; Kim and Ahn, 2022).

East Africa like other parts of the world face the burden of AMR, despite this burden still there is limited studies on AMR including gene distribution and population structure. The disease determinants in East Africa for *C. jejuni* AMR including but not limited to limited stewardship programs, misuse and overuse of antibiotics in animals and human, poor sanitation and weak veterinary and health infrastructure (Kaakoush et al.,2015; Tang et al., 2017). Studies have reported high ciprofloxacin resistance attributed by *C. jejuni* in Kenya and Tanzania (Mshana et al., 2016) and azithromycin resistance by *C. jejuni* genes in Uganda (Kateete et al., 2019).

The proper way of addressing AMR for *C. jejuni* requires a global and regional joint effort (experts and rapid diagnostic tools) and integration of one health approach (human, animal, and environmental surveillance), as well as active genomic surveillance through the use of WGS. WGS plays a significant role in AMR by tracking and identifying resistant genes, transmission ways, and genetic diversity in bacterial strains (Cody et al., 2012). WGS detects *C. jejuni* resistant genes like gyrA (Fluoroquinolone resistant) and ermB, 23S rRNA (Macrolides resistant) (Luangtongkum et al., 2009). Other genes, such as tet(O) conferring tetracycline resistance and cmeABC efflux pump systems contributing to multidrug resistance, have also been widely reported (Wieczorek & Osek, 2013; Duarte et al., 2014).

Moreover, integration of the WGS and bioinformatics is important for bridging critical knowledge gap in AMR management. These techniques help to improve surveillance beyond simple monitoring to powerful, evidence-based system that clarifies the complex ecology of resistant pathogens including *C. jejuni* (WHO, 2021). Through provision of high-resolution insights in evolutionary dynamics and transmission pathways at human-animal and environmental interface (WHO, 2021). Additionally, they provide data necessary to design interventions such as antibiotic stewardship programs and biosecurity measures, which in turn help mitigate the global spread of AMR (WHO, 2021). Furthermore, WGS-guided risk assessment can inform policymakers on the effectiveness of control strategies in food production, such as improved farm hygiene, reduced prophylactic use of antibiotics in livestock, and vaccination research (EFSA & ECDC, 2021; Tang et al., 2017). This integrated strategy contributes to Sustainable Development Goal 3, which aims to ensure healthy lives and promote well-being by reducing infectious diseases and drug resistance (United Nations, 2015).

The key resistant mechanism in *C. jejuni* have been tabulated in table 1 below.

**Table 1.** Resistance mechanism of *C. jejuni*.

| Antibiotic class | Resistant genes | Resistance mechanism | References |
| --- | --- | --- | --- |
| Fluoroquinolones | *gyrA* mutations (Thr-86-Ile) | Altered DNA gyrase target | (Wieczorek et al., 2018) |
| Macrolides | *ermB*, *23S rRNA* mutations | Ribosomal modification | (Tang et al., 2017) |
| Tetracyclines | *tet(O)* | Ribosomal protection | (Iovine, 2013) |
| Beta-lactams | *blaOXA-61* | Beta-lactamase production | (Alfredson & Korolik, 2007) |

This study applies a One Health Approach and genomic surveillance to analyze WGS data from Ethiopia, Kenya and Tanzania in East Africa, with a primary focus on *C. jejuni* circulating among humans, animals and environment. Using WGS, the study aims to address existing knowledge gaps by characterizing AMR gene profiles, virulence determinants, and the genomic diversity of *C. jejuni* isolated from *Homo sapiens, Gallus gallus, Bos indicus, Ovis aries*, goat and milk from dairy cattle. The findings will provide genomic evidence to strengthen surveillance systems, support antimicrobial stewardship and advance One Health strategies to address the growing global challenge of AMR.

## METHODS

### Data acquisition

A comprehensive literature search was conducted across various scientific journals and databases using key terms such as “whole genome of *Campylobacter jejuni* in Tanzania”, “whole genome of Campylobacter in Africa”, “whole genome of campylobacter in sub-Saharan” and “antibiotic resistance in livestock, human and environment”.

Relevant studies were screened to identify and obtain corresponding project and accession number(s) which were subsequently used to obtain the whole genome sequences (WGS) data from public repositories for analysis.

All raw WGS data were retrieved from the NCBI GenBank database by using SRA-Toolkit through Ubuntu command line interface based on the project and accession numbers listed in table 2.

**Table 2.** List of accession numbers for genomic sequences included in this study, the accession numbers correspond to *C. jejuni* isolates analyzed across various sources including *Homo sapiens, Gallus gallus, Bos indicus, Ovis aries,* goat and milk from dairy cattle.

| SN | Accession number | Source of sequence | Number of sequences | Reference |
| --- | --- | --- | --- | --- |
| 1 | PRJNA1015272 | NCBI-SRA | 64 | (Bardosh et al., 2020) |
| 2 | PRJNA1026168 | NCBI-SRA | 83 | (French et al., 2024) |
| 3 | PRJNA578397 | NCBI-SRA | 14 | (Admasie et al., 2024) |

### Data processing (Quality control)

Fastqc v0.12.1 was used to assess the quality of raw reads. The retrieved reads were all of good quality with the quality score (Q) greater than 20; thus, there was no need for trimming since quality scores were greater than 20 which is within tolerable limit for Illumina data, hence cannot interfere with subsequent downstream analysis (Omar et al., 2024).

### Genome assembly

Genome Assembly was performed by using SPAdes (Bankevich et al., 2012), where paired-end reads were assembled into contiguous sequences. Prokka software was used for genome annotation (Seeman, 2014). Single Nucleotide Polymorphism (SNP) variant calling was done using bcftools software (Danecek et al, 2021). The workflow involved mapping sequences to the *C. jejuni* reference genome (*C. jejuni* subsp. jejuni NCTC 11168 = ATCC 700819), then SAM files were generated which in turn converted to BAM files and mpileup was generated. The bcftools was used to generate VCF files for each sample and then individual VCF files were merges and single VCF file was formed using bcftools.

### AMR gene identification

Contig FASTA sequences generated by SPAdes during genome assembly were used to identify the presence of AMR genes using ResFinder software (Zankari, 2014). In-house Python script, followed by the heatmap in R was employed to create heatmap visualization. Then the ggVennDiagram function in R was used to illustrate the presence and distribution of the shared and unique (ARGs) across three different sources and three different countries.

### Virulence gene identification

The contigs.fasta sequences produced by SPAdes software for each paired end read were analyzed for *C. jejuni* virulence factors. Virulence genes in all contigs.fasta sequences were identified using Abricate software (Seeman, 2020) with the virulence factor database (VFDB) (Liu et al., 2022). In-house Python script was used to generate a heatmap visualization of the virulence-associated genes.

### Population structure

Wright Fixation Index (FST) and Principal component analysis (PCA) were used to access gene flow among *C. jejuni* isolates from *Homo sapiens*, *Gallus gallus*, milk (raw and pasteurized from dairy cattle), goats, *Bos indicus* and *Ovis aries* (Danecek et al., 2021) and population structure was determined using principal component analysis (PCA) as implemented in PLINK 1.9 (van Waaij, 2023).

### Analysis of the genetic similarities among Antimicrobial -Resistant strains

Multilocus Sequence Typing (MLST) was used to access the genetic relatedness among anti-microbial resistant isolates. MLST version 2.23.0 was implemented in Linux environment and the resulting data were visualized in R using the ggraph and tidygraph packages to illustrate the genetic connectedness between isolates.

## RESULTS

### AMR genes identification and distribution

A total of 161 WGS sequences were obtained from the NCBI database and analyzed, (130) results showed the distribution of AMR genes across seven (7) key sources with seven (7) resistance genes found in multiple sources (Table 3). *ARGs conferring resistance to* β-lactams, tetracyclines and aminoglycosides were identified from *Homo sapiens*, Milk (from dairy cattle) and *Gallus gallus.* Remarkably blaOXA-193 was present across all three countries, tetracycline was present in Ethiopia and Tanzania mainly in *Gallus gallus* and milk (from dairy cattle), Kenya was limited to β-lactamases exclusively in *Homo sapiens*.

**Table 3.** The distribution of (ARGs) across Ethiopia, Kenya and Tanzania, ARGs were grouped according to antimicrobial classes and sources of isolation.

| Country | ARG | Class of antimicrobials | Source of organism |
| --- | --- | --- | --- |
| ETHIOPIA | blaOXA-193 | beta-lactams | Pasteurized milk (n=3), raw milk (n=12), *Homo sapiens* (n=28), *Gallus gallus* (n=27), Goat (n=2), *SRR26351312, *Ovis aries* (n=2), *Bos indicus* cow (n=1) |
|  | tet(O) | tetracycline | Pasteurized milk (n=2), Raw milk (n=7), *Homo sapiens* (n=5), *Gallus gallus* (n=2) |
|  | tet(O/32/O) | tetracycline | *Homo sapiens* (n=7), *Gallus gallus* (2) |
|  | ant (6)-Ia_3 | tetracycline | *Gallus gallus* (n=1) |
| TANZANIA | blaOXA-193 | beta-lactams | Pasteurized milk (n=3), Raw milk (n=12), *Gallus gallus* (n=17) |
|  | tet(O) | tetracycline | Pasteurized milk (n=2), Raw milk (n=7), *Homo sapiens* (n=2), *Gallus gallus* (n=20) |
|  | tet(O/32/O) | tetracycline | *Gallus gallus* (n=3) |
|  | ant (6)-Ia_3 | aminoglycoside | *Gallus gallus* (n=2) |
|  | blaOXA-489 | beta-lactams | *Gallus gallus* (n=2) |
| KENYA | blaOXA-193 | beta-lactams | *Homo sapiens* (n=19) |
|  | blaOXA-489 | beta-lactams | *Homo sapiens* (n=2) |
|  | blaOXA-61 | beta-lactams | *Homo sapiens* (n=2) |
|  | blaOXA-184 | beta-lactams | *Homo sapiens*(n=1) |

n=Number of samples
\*= Unknown

### Heatmap and hierarchical clustering of AMR profiles

The heatmap shows presence and absence of AMR genes found across 161 assembly results. Red color (value=1) indicates presence of AMR genes, blue color (value =0) indicates absence of AMR genes. The hierarchical clustering shown by the dendrogram at the left and top side of the heatmap have grouped samples (accession numbers) and genes based on the similarity of the AMR genes found (profiles).

AMR genes such as blaOXA-193, tet(O), tet(O/32/O) and ant (6)-Ia were found across all assemblies, this indicates the presence of limited AMR genes diversity. On other hand majority of other genes were absent as indicated by large blue region in the heatmap below. Moreover, samples with shared AMR genes clustered together indicating presence of shared resistance profiles, this have shown from Isolates harboring blaOXA-type β-lactamases such as blaOXA-193, blaOXA-489, blaOXA-61, blaOXA-184 and another group was tetracycline resistant genes tet(O) clustered together respectively.

Therefore, heatmap show that there is high prevalence of β-lactamases genes due to detection of several blaOXA variants across isolates, this reflect widespread of β-lactam resistance which is common in *C. jejuni* and other gram-negative bacteria mainly associated with intrinsic or acquired resistance. On other hand tetracycline resistance tet(O) genes appears prominently suggesting frequent resistance to tetracyclines and few isolates shown ant (6)-Ia gene presence but it is much less frequent compared to β-lactam and tetracycline genes indicating low distribution of the aminoglycoside resistance.

### Shared and Unique AMR genes among the host

In examination of the overlap and difference in distribution of resistance genes between host species a Venn diagram was generated (Figure 3). This analysis illustrates unique and shared antimicrobial resistance genes among *Homo sapiens, Gallus gallus* and livestock indicating the extent of overlap across hosts. The majority of ARGs were predominantly associated with *Gallus gallus*, 41.7% and *Homo sapiens*, 33.3%. *Homo sapiens* and *Gallus gallus* shared a smaller proportion of genes (16.7%). Only one gene (8.3%) was shared between *Homo sapiens* and *Ovis aries*.

**Figure 1.**
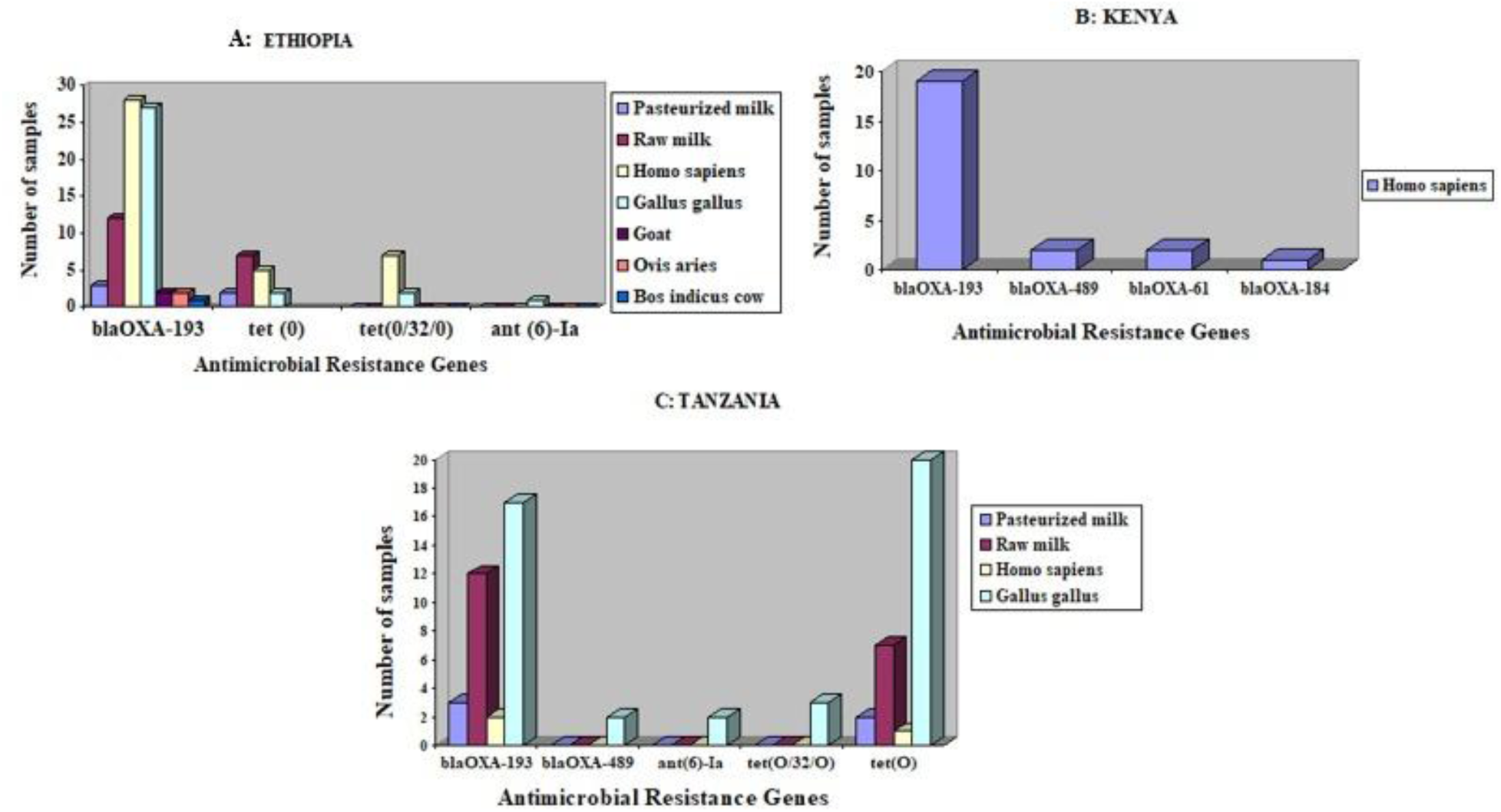
(A-C) Illustrates the distribution of AMR genes in *C. jejuni* from Ethiopia, Kenya and Tanzania respectively. The most widespread gene was blaOXA family, blaOXA-193 was predominantly spread in Ethiopia and Tanzania and also observed in Kenya *Homo sapiens* isolates together with blaOXA-489, blaOXA-61, blaOXA-184. Tetracycline genes (tet(O) and tet(O/32/O) were most common in Ethiopia and Tanzania and aminoglycoside genes (ant(6)-Ia was identified only in Ethiopia and Tanzania poultry.

**Figure 2.**
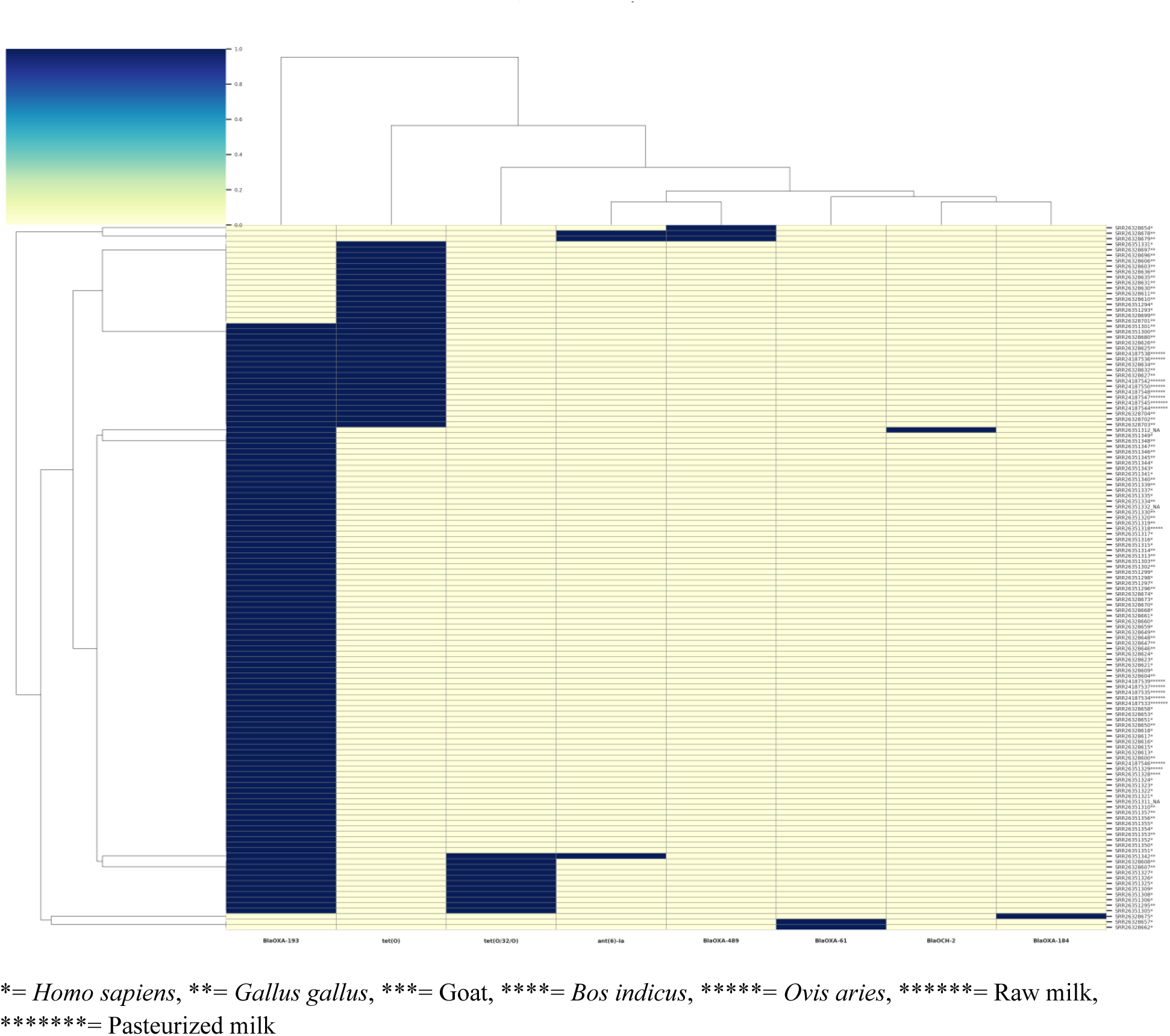
Presence or absence heatmap of antimicrobial resistance genes in *Campylobacter jejuni* isolates from Ethiopia, Kenya and Tanzania. Genes were shown on the x-axis and the isolates were clustered on the y-axis with host and source annotation as described in the key below the heat map. The gradient scale display absence (0.0) to presence (1.0) of each gene. blaOXA-193 was remarkably to be the common across all hosts and sources, confirming it as the main component of the regional resistome. On other hand tetracycline (tet(O) and tet(O/32/O) were detected in *Homo sapiens*, *Gallus gallus* and dairy sources, blaOXA-489 appeared mainly in Tanzania *Gallus gallus*, blaOXA-61 and blaOXA-184 occurred only in Kenya *Homo sapiens* isolates and aminoglycosides ant(6)-Ia was limited to *Gallus gallus* isolates.

**Figure 3:**
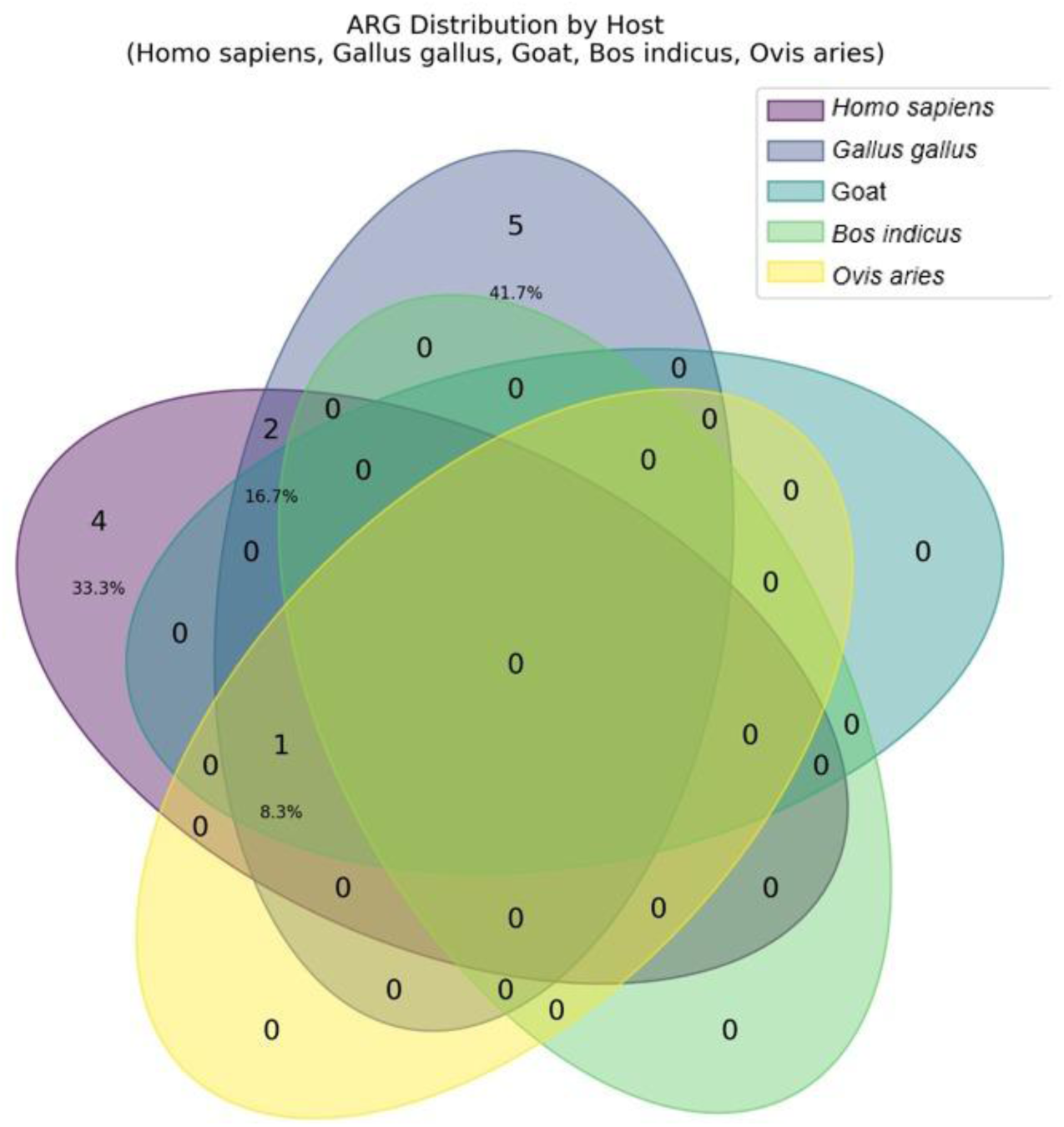
Venn diagram displaying the distribution of AMR genes among different hosts which include *Homo sapiens*, goats, *Bos indicus*, *Ovis aries* and *Gallus gallus*.

### Distribution of AMR across Countries

In examination of the geographic variation, AMR determinants were stratified based on origin (Ethiopia, Tanzania and Kenya). Relative frequency of key AMR genes identified including blaOXA variants (blaOXA-193, blaOXA-489, blaOXA-61, blaOXA-184), tetracycline determinants (tet(O) and tet(O/32/O) and aminoglycoside makers were summarized (Figure 4).

**Figure 4.**
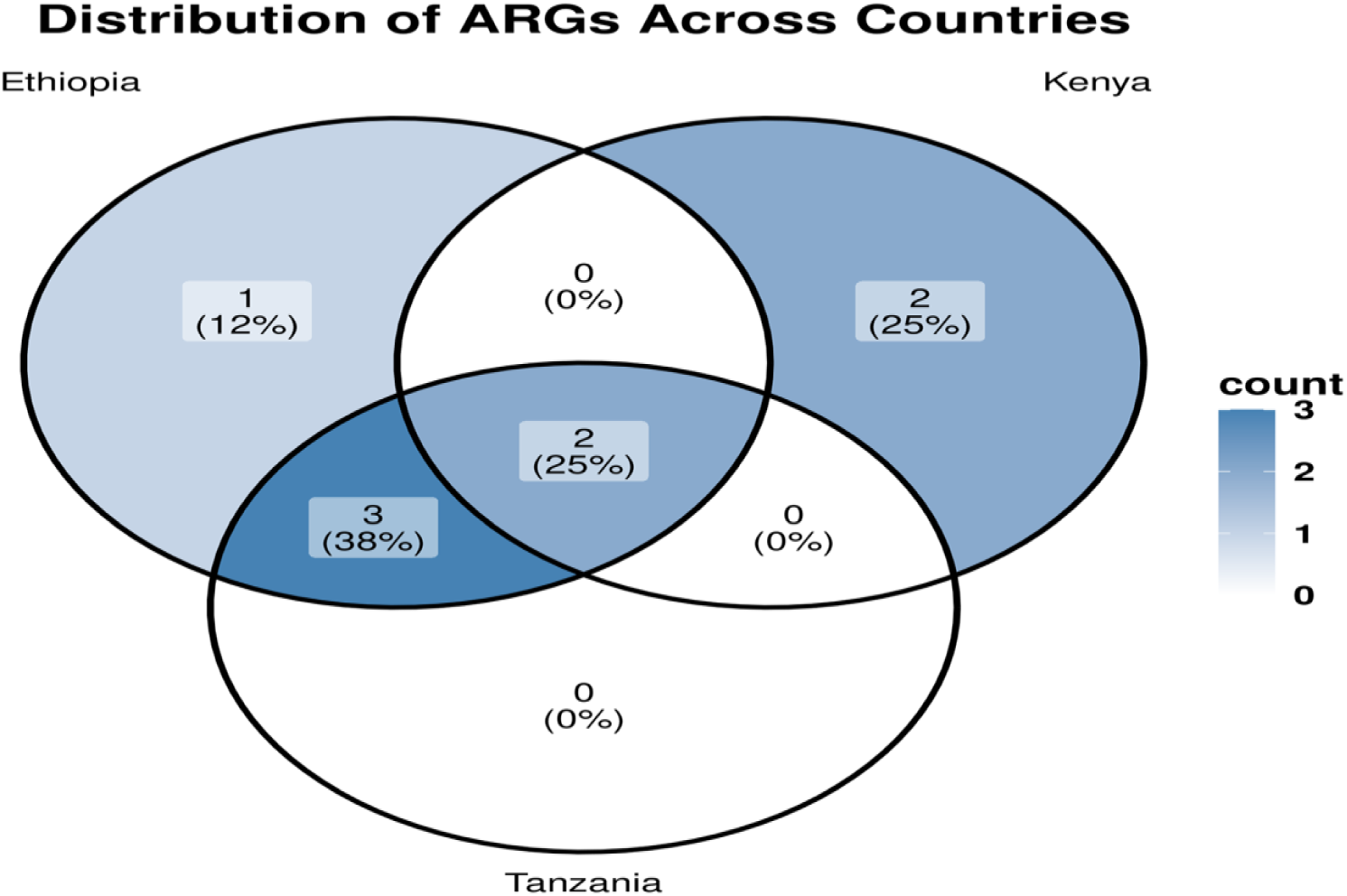
Venn diagram illustrating the distribution of the (ARGs) in Ethiopia, Kenya and Tanzania. One gene (12%) of the total ARGs was unique in Ethiopia, 25% of total ARGs were unique in Kenya. Three ARGs (38%) were common in Ethiopia and Tanzania, and two ARGs (25%) were shared to all three countries and no ARGs were unique to Tanzania.

This illustrate that Ethiopia shows presence of broadest AMR genes profile, followed by Kenya including rare blaOXA alleles (blaOXA-184/-489) primarily among *Homo sapiens* isolates and Tanzania is dominated by the blaOXA-193 with wide spread tet(O). Therefore, these patterns display presence of shared cross border resistome with country specific edges likely shaped by local antimicrobial us e and value chain practices.

### Virulence genes identified

The prevalence of virulence-associated genes across all 161 genomes were summarized in Table 4 below and genes were categorized into very high prevalent (>140), moderately high prevalent (120-140), moderately (100-138), moderately low prevalent (50-100), low prevalent (<50) and very low (rare markers) prevalent (≤1). This result highlights the dominance of adhesion and motility genes such as flg genes, cadF and fli genes, cytolethal distending toxin genes such as cdtA, cdtB, and cdtC. Furthermore, presence of capsule biosynthesis loci responsible for bacteria protection and pathogenicity such as kps genes were also observed. This result shows both conserved core virulence determinants and less common genes which is due to strain-specific pathogenicity

**Table 4.**
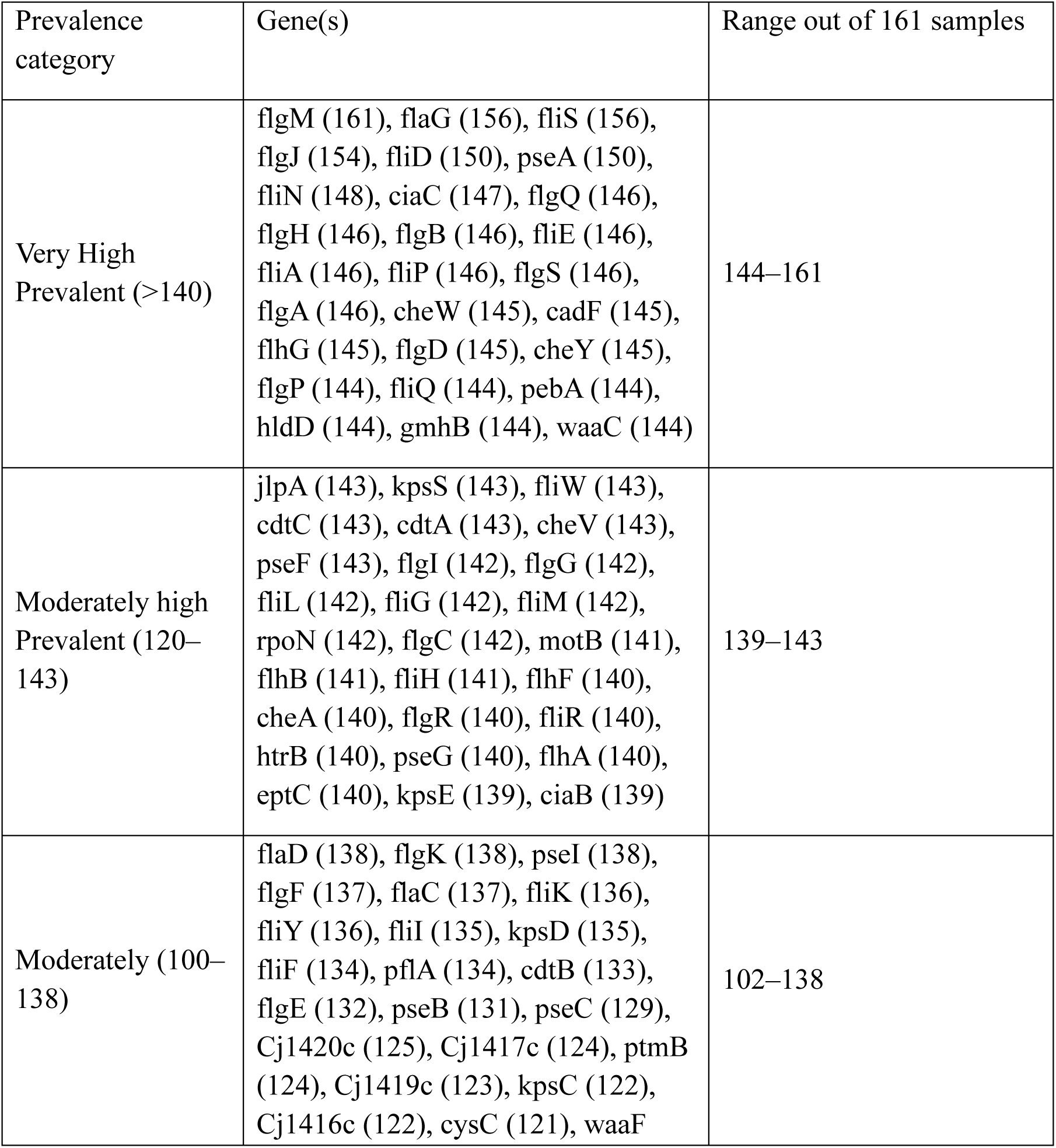
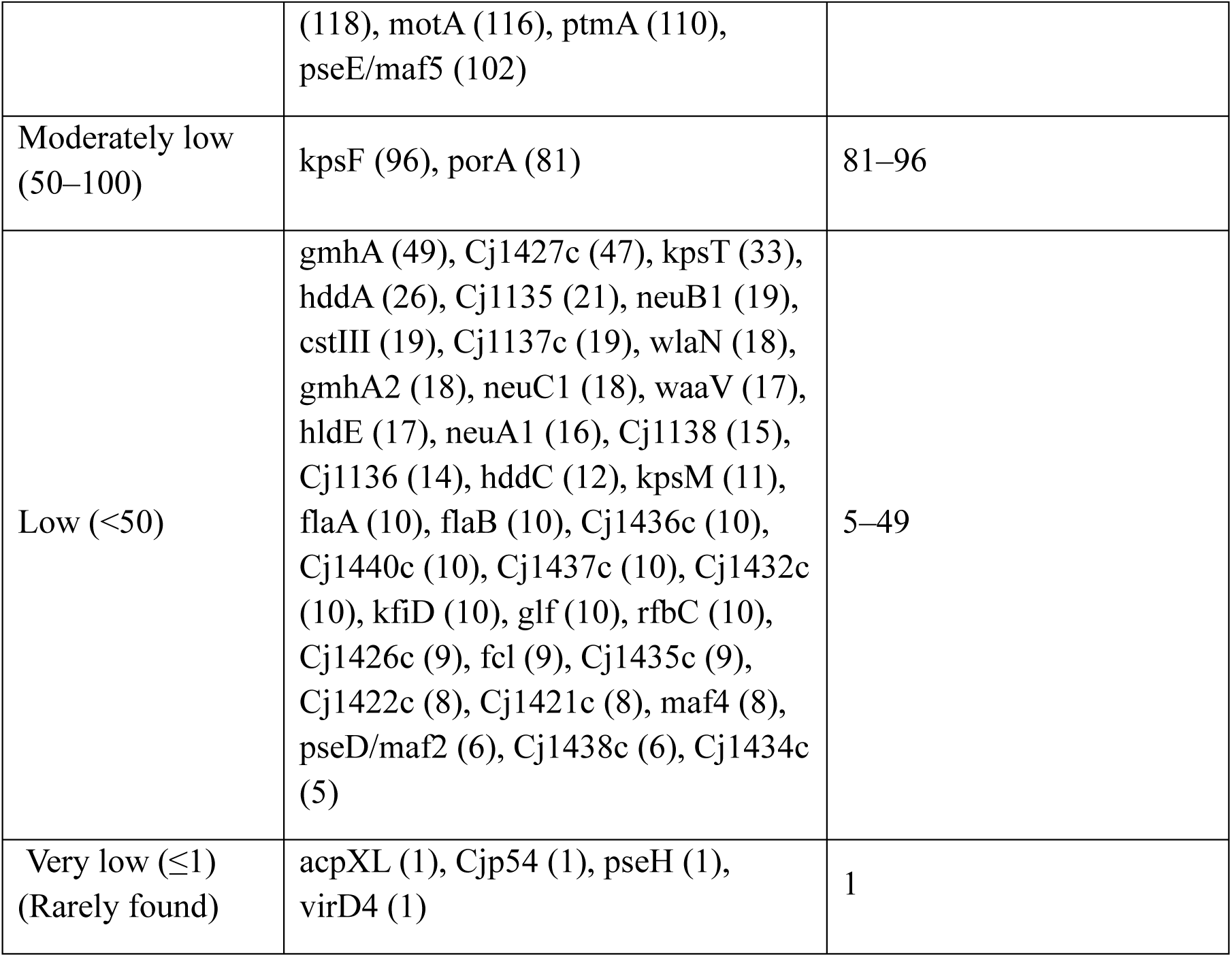
The prevalence of virulence-associated genes across all 161 genomes. genes were categorized into very high prevalent (>140), moderately high prevalent (120-1420), moderately (100-138), moderately low prevalent (50-100), low prevalent (<50) and very low (rare markers) prevalent (≤1).

To further visualize pathogenic profile and complement table 4 above, the frequency and distribution of obtained virulence genes were displayed in heatmap highlighting conserved virulence genes profiles across isolates as seen in figure 5 below. Each row represents an isolate (accession number and host species) and each column represent the virulence genes determinants identified. Hierarchical clustering of isolates was displayed in dendrogram based on the virulence genes profile. This heat map reveals that a highly conserved virulence genes profiles was dominated by adhesion genes (cadF, jlpA), motility and flagellar genes (fli, flg families), and the cytolethal distending toxin cluster (cdtA, cdtB, cdtC), which were consistently present across nearly all isolates. Capsule biosynthesis loci (kps genes) as well as other pathogenic factors were also wide spread while other genes were appeared sporadically.

**Figure 5.**
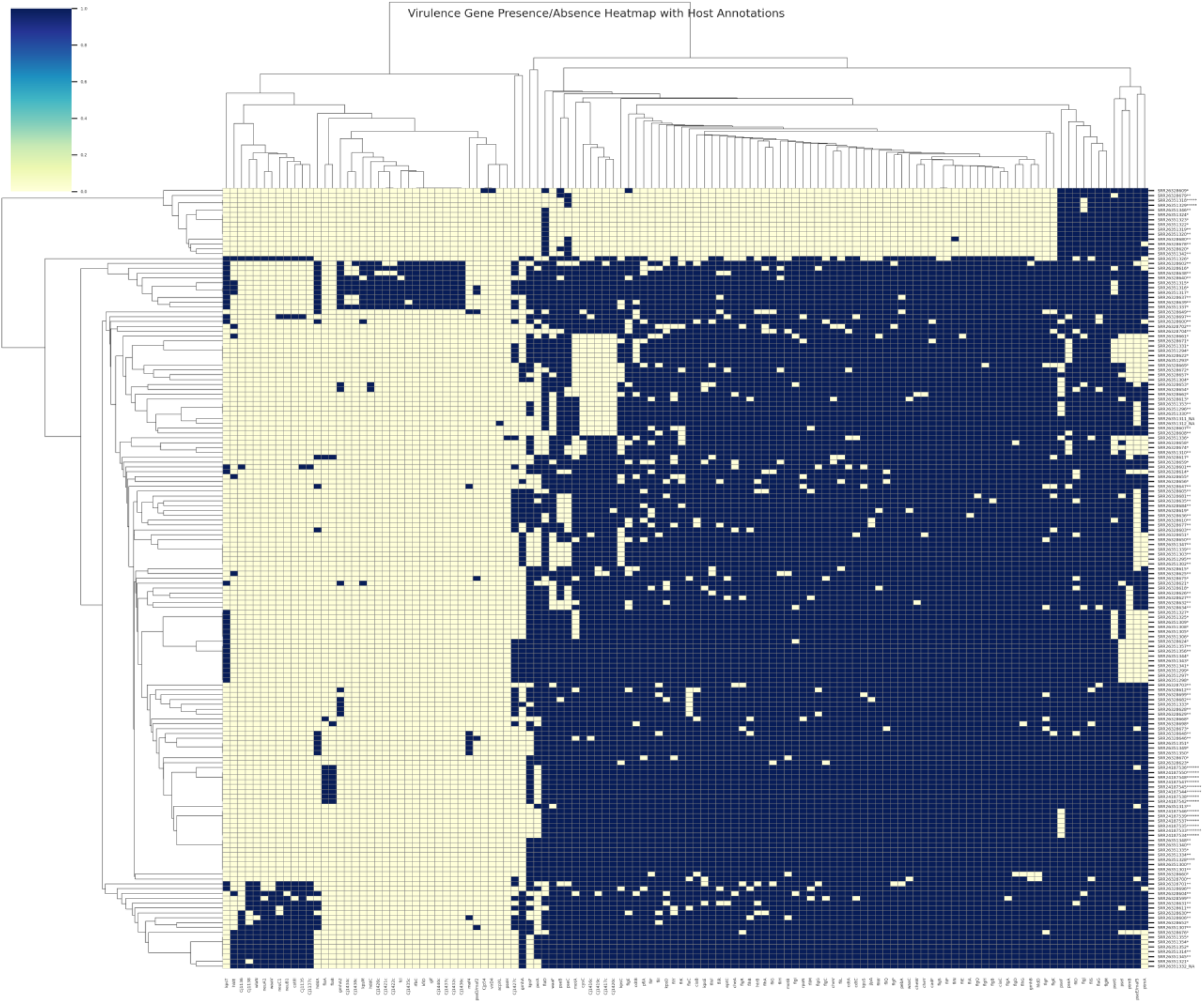
Heatmap of presence/absence with host annotation for 161 *C. jejuni* isolates from Ethiopia, Kenya and Tanzania showing the presence (dark blue) or absence (light yellow) of virulence linked genes across genomes. Virulence genes were shown on the x-axis and the isolates were clustered on the y-axis.

### Population structure

This section explained how the population structure of *C. jejuni* isolates was assessed using (PCA) based on SNPs data. The PCA results showed that clustering by host and geography occurred, but there was also frequent overlap between *Gallus gallus* and *Homo sapiens* isolates, indicating close genetic relatedness. When all host categories *Homo sapiens*, *Gallus gallus, Bos indicus, Ovis aries,* goat, and milk (from dairy cattle), were included (Figure 6), the isolates demonstrated strong intermixing, which supported evidence of cross-host transmission pathways.

**Figure 6.**
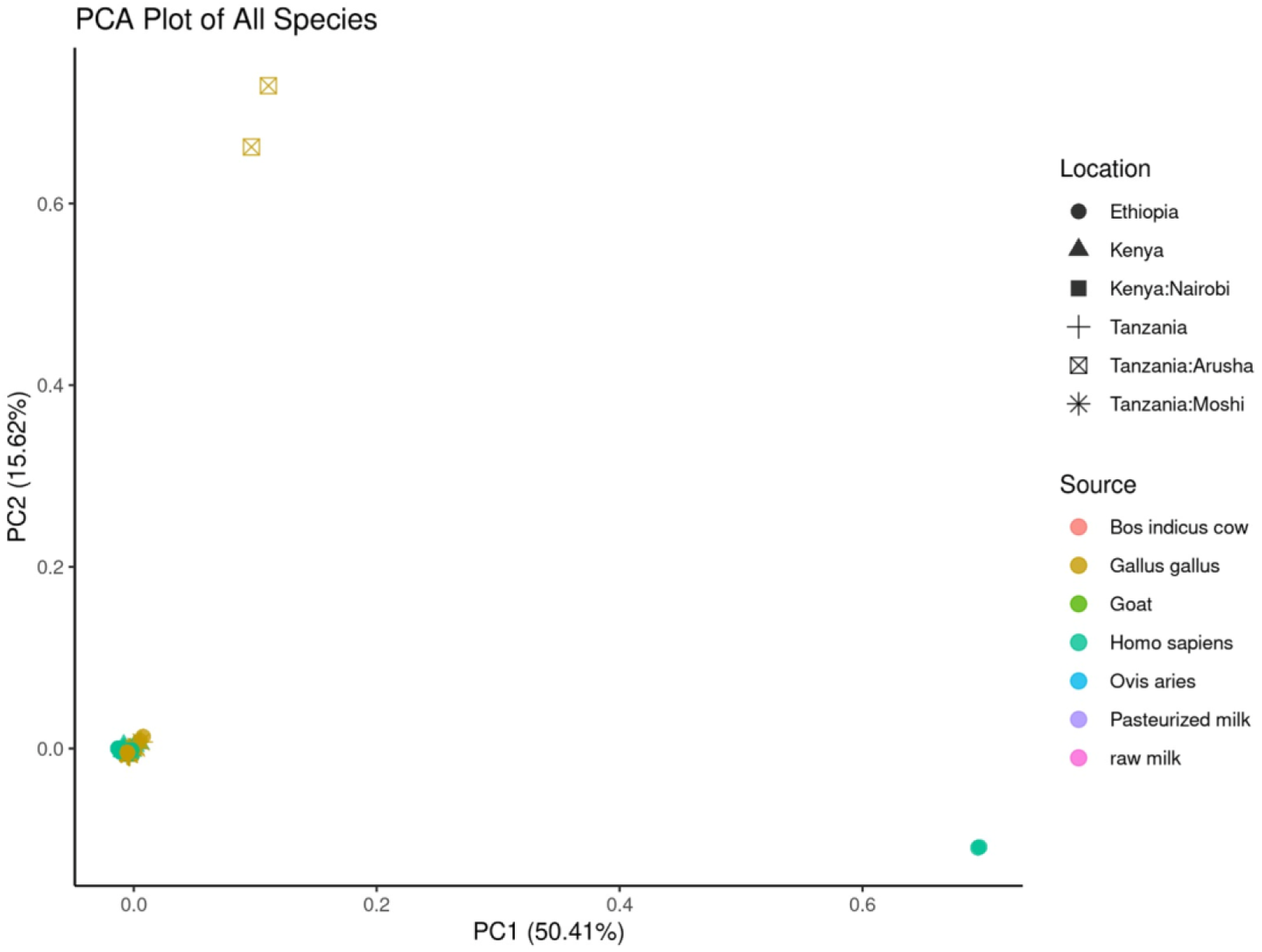
Principal Component Analysis (PCA) plot displaying the genetic spreading of *C. jejuni* isolated across different sources and geographic location. It confirms the overall clustering patterns indicating limited genetic differentiation among isolates from various hosts and regions.

To contextualize the dataset composition, isolates originating from each source was summarized. The pie chart (Figure 7) illustrates the dominance of *Homo sapiens* and *Gallus gallus* derived isolates with smaller contributions from milk, goats, *Bos indicus* and *Ovis aries*.

**Figure 7.**
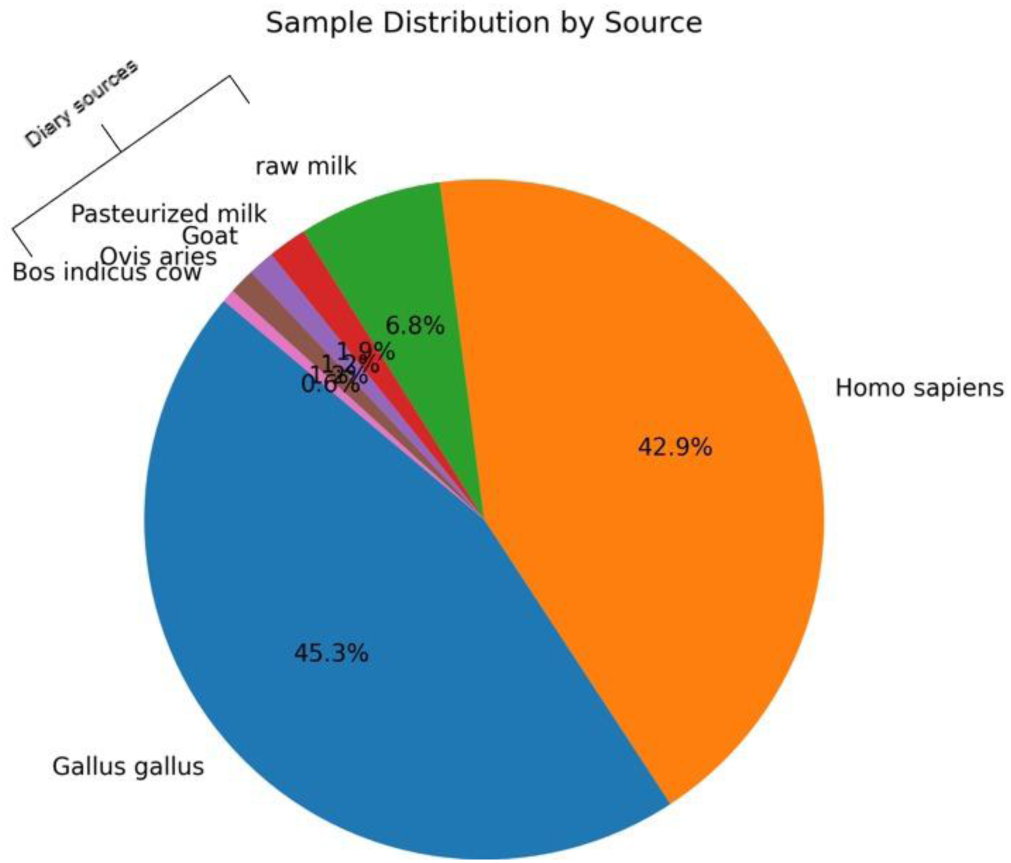
A pie chart shows sample contribution by source in which *Homo sapiens* and *Gallus gallus* contributed the greatest quantity, making up 42.9% and 45.3% of the total samples, respectively. Pasteurized milk made up 1.9%, and raw milk made up 6.8%. Goats accounted for 1.2% of the total, *Ovis aries* for 1.2%, and *Bos indicus* for 0.6%. Nearly 90% of the isolates were from *Homo sapiens* and *Gallus gallus* sources combined, highlighting their crucial importance in the dataset in contrast to the comparatively little contributions from dairy and ruminant sources (11.8%).

### Minnimum spanning tree (MST)

Minnimum spanning tree was constructed to evaluate genetic relatedness among isolates. The MST (Figure 11) display denser clustering of *Homo sapiens* and *Gallus gallus* isolates with scattered milk, goat, *Ovis aries* isolates suggesting and confirming strong connectivity across hosts.

## DISCUSSION

This study applied a One Health genomic surveillance framework to examine antimicrobial resistance (AMR), virulence, and population structure of *C. jejuni* isolates from Ethiopia, Kenya and Tanzania. Whole-genome sequencing (WGS) revealed a restricted but clinically important resistome dominated by β-lactamases (blaOXA family) and tetracycline determinants (tet(O), tet(O/32/O), with limited aminoglycoside genes and rare macrolide determinants. Results from Table 3 and figure 1(A-C) give an overview of the detected ARGs and the possible Multi-Drug Resistant (MDR) genes from human, animals and food samples across three countries, based on WGS analysis. Among the ARGs β-lactam (blaOXA) genes which confer resistance to β-lactam drugs such as ampicillin, penicillin and amoxicillin were found to be predominant in all countries. Specifically, blaOXA_193 and blaOXA_61 genes were widespread in *Homo sapiens* (human) and followed with *Gallus gallus* (poultry), small ruminants (goat, *Ovis aries*, *Bos indicus*) as well as in raw and pasteurized milk. This indicates that the resistome of blaOXA_193 gene is highly conserved in *C. jejuni* in region under study. Habib et al., 2023 study from UAE found poultry to be highly dominated by blaOXA_193 and blaOXA-61 resistance genes. The blaOXA184 was found in human sample from Kenya while blaOXA489 were found in poultry and human sample from Tanzania and Kenya respectively. The tet(O) and its invariant tet(O/32/O) genes were found in human, food and animal samples with Tanzania being higher than Ethiopia indicate the high chance of having resistance to tetracyclines antibiotics due to frequently being used for bacterial infection treatment and growth promotion in veterinary medicine for across East African countries (Iovine, 2013; Kimera et al.’ 2020). Occasionally ant (6)-Ia gene conferring resistance to aminoglycoside class of antibiotics was found in *Gallus gallus* sample across Tanzania and Ethiopia.

The distribution of resistance genes across human, animals and food indicate the possible horizontal gene transfer across the organisms through environmental contamination or farm to fork chain. These results align with global findings that *C. jejuni* maintains a relatively narrow but stable set of resistance mechanisms, often horizontally transferred (Wieczorek et al., 2018; French et al., 2024).

The AMR heatmap (figure 2) demonstrate that most isolates harbored one or two resistant determinants, dominated by blaOXA alleles (blaOXA variants (blaOXA-193, blaOXA-489, blaOXA-61, blaOXA-184) and tetracycline determinants (tet(O)). On the other hand, clustering discovered the unique β-lactamase-dominant and tetracycline-dominant lineages, with limited evidence for broad multidrug resistance. Analysis of Venn diagram (Figure 3) displayed the overlap of the blaOXA-193 and tet(O) between *Homo sapiens* and *Gallus gallus* isolates, highlighting zoonotic transmission. Country wise Ethiopia has broad gene diversity followed by Kenya showed rare alleles such as blaOXA-184/-489 while on other hand Tanzania was dominated by blaOXA-193 and tet(O). The findings of this study concur with Admasie et al., (2024) who reported dominance of blaOXA and tet(O) in Ethiopia’s dairy chain and Frenc et al., (2024) who reported the overlapping *C. jejuni* STs and AMR genes between human and poultry isolates in East Africa.

Analysis revealed that *C. jejuni* isolates carried a conserved set of virulence genes, including adhesion markers such as cadF, jlpA, motility and flagellar genes such as flg, fli families and the cytolethal distending toxin cluster such as cdtA, cdtB and cdt C as seen in Table 4 and visualized in Figure 5. Capsule biosynthesis genes such as kps and waa were also observed in isolates. This resembles global datasets showing cadF, flagellar genes and cdt families as part of the essential core virulome (Panzenhagen et al., 2021). Similarly to East Africa a European WGS study also found that Italian *C. jejuni* carried abundant virulence markers, (Garcia-Fernandez et al., 2024) as presented in our results.

Regional comparisons strengthen these observations. In Kenya, cadF, flaA, ciaB, and cdtA were among the most common virulence markers, with cadF and flaA strongly correlated (Wanja et al., 2024). Tunisian poultry isolates frequently carried cadF and ciaB, and these were significantly associated with multidrug resistance, suggesting synergy between AMR and virulence traits (Gharbi et al., 2022). In Brazil, poultry isolates displayed higher frequencies of flaA and cdtA compared with human isolates, supporting poultry as a reservoir of highly virulent strains (Lima et al., 2022). In Chile, cdtA,cdtB,cdtC and capsule loci were widespread, and additional Type VI and Type IV secretion systems were reported, expanding the virulence landscape of South American isolates (Bravo et al., 2021).

Clinical studies underline the pathogenic importance of these determinants. In Bahrain, cdtB and iam were significantly associated with symptomatic infections, particularly in young children (Al-Mahmeed et al., 2006). The high prevalence of cdtA, cdtB, and cdtC in the analyzed isolates aligns with evidence linking CDT toxins to diarrheal disease severity.

Furthermore, East African *C. jejuni* isolates carry virulence markers relative to some other global datasets that are potential to their capacity for adhesion, invasion, and toxin-mediated pathogenesis. These findings of β-lactamase resistance genes with strong virulence backbone raise a concern for both clinical management and One Health surveillance.

The PCA provided important insights into the genetic relatedness and population structure of *C. jejuni* isolates across different hosts and geographic regions. The clustering patterns that were observed demonstrated both host- and geography-associated structure; however, the frequence overlap of *Gallus gallus* and *Homo sapiens* isolates indicated that *Gallus gallus* remained a primary reservoir for human infection. This finding aligned with previous studies that highlighted poultry as the dominant source of human campylobacteriosis worldwide (Sheppard et al., 2014; Cody et al., 2012).

Importantly, the broader PCA incorporating all host categories revealed substantial intermixing of isolates. This finding supported the hypothesis of multi-host transmission dynamics, where *C. jejuni* strains were capable of infecting a wide range of reservoirs, including livestock, milk, and the environment, in addition to poultry and humans. The intermixing of isolates across sources underscored the complexity of transmission pathways and highlighted the role of indirect exposure routes such as contaminated food, water, and animal products (Elbehiry et al., 2025).

The proportional analysis further contextualized the dataset composition, showing the predominance of poultry- and human-derived isolates. This distribution reflected both the epidemiological importance of poultry as a major infection source and potential sampling biases toward human clinical and poultry isolates, which were more accessible than those from other reservoirs. Nevertheless, the smaller but significant contributions from cattle, sheep, goats, and milk highlighted the potential for secondary reservoirs to act as sources of spillover infections.

Collectively, these findings reinforced the need for a One Health approach in controlling *C. jejuni* infections. The evidence of genetic overlap across hosts and regions emphasized that interventions targeting a single host or setting were unlikely to be sufficient. Instead, integrated surveillance across human, animal, and environmental compartments, coupled with genomic monitoring, was essential for identifying transmission hotspots and informing risk-based control strategies. Furthermore, the observed intermixing of isolates highlighted the necessity of strengthening biosecurity measures in poultry production and improving food safety interventions throughout the supply chain to reduce cross-host and zoonotic transmission risks.

The MST (Figure 8) demonstrated genetic overlap between poultry and human isolates while other isolates like milk, goat, sheep and cow interspersed, this is important since it depict that poultry as the dominant reservoir with strong emphasis of the cross-species spillover potentials. The findings of this study align with East Africa WGS survey showing extensive overlap between poultry and human for *C. jejuni* (French et al., 2024) and support the One Health perspective.

**Figure 8:**
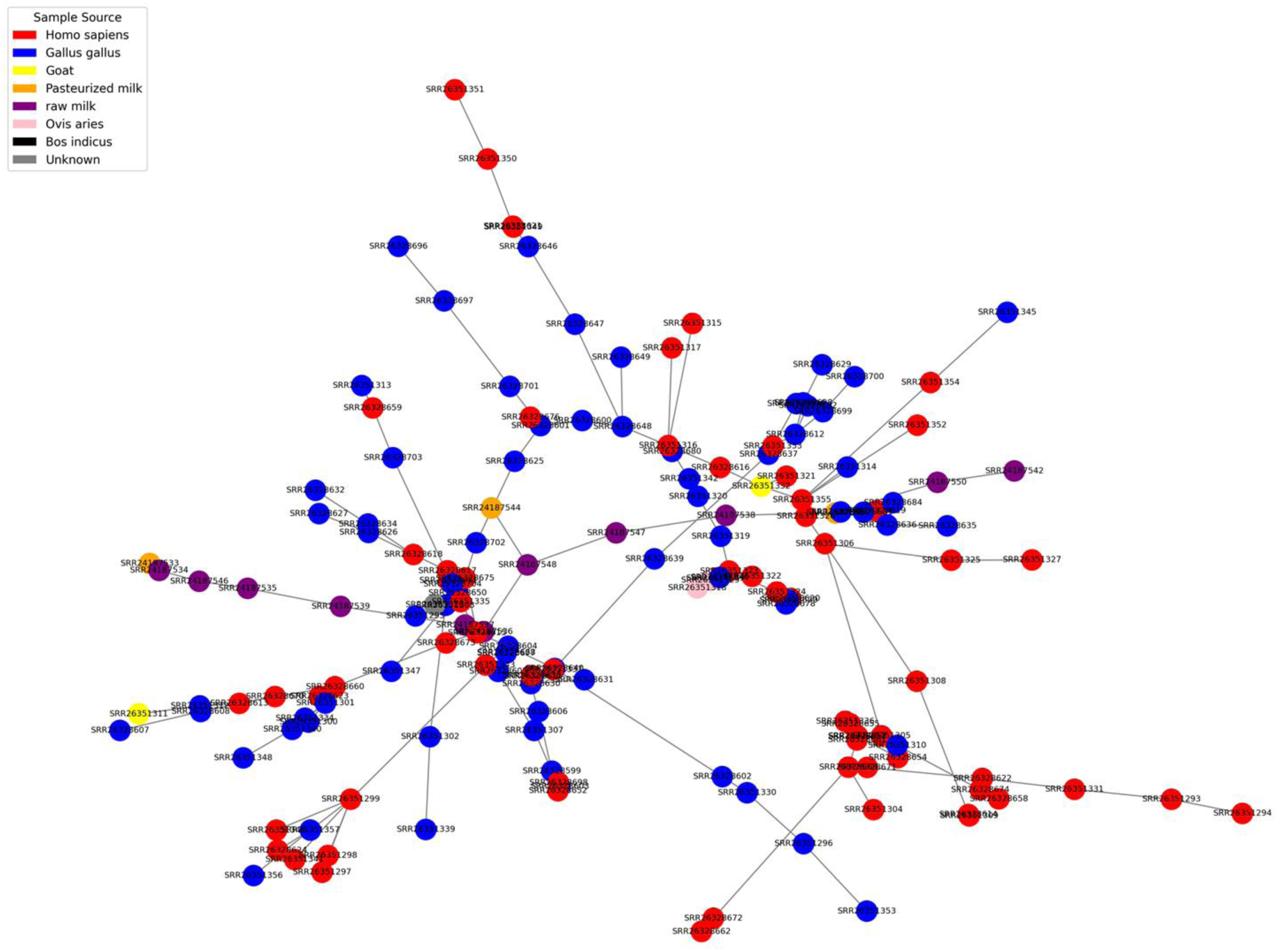
Minimum spanning tree based on core genome Multilocus Sequence Typing (cgMLST). Each node represents an isolate (SRR accession number) with color coded by sample sources and the edge represent the relatedness. The branches represent potential evolutionary trajectories.

### Conclusion and Recommendations

This study demonstrates that *C. jejuni* in Ethiopia, Kenya, and Tanzania carry a restricted but important resistome dominated by blaOXA β-lactamases and tetracycline resistance genes, shared across humans, poultry, and dairy sources. Rare blaOXA variants (blaOXA-489 and blaOXA-184) in Kenya add to regional diversity, while virulence profiling shows a conserved pathogenic backbone with adhesion, motility, and toxin genes. Poultry emerged as the dominant reservoir and plasmid replicon analysis suggested possible horizontal transfer of AMR genes across species.

Therefore, it’s important to strengthened antibiotic stewardship, invest more in sustainable genomic surveillance, diagnostics and biosecurity further integrate the scientific evidence results in National AMR strategic plan and national guidelines for drug use schedules. In consideration to this study for campylobacteria’s macrolides and fluoroquinolones need to be preserved for effective use in future. Further studies should also focus more on comparative ARG modelling and transmission dynamic across the three countries

### Implications in one health

The findings confirm poultry and humans as key reservoirs of AMR genes in *C. jejuni*. The persistence of blaOXA variants threatens penicillin efficacy, while widespread tetracycline resistance challenges veterinary treatment. The low prevalence of macrolide and fluoroquinolone resistance offers a window to preserve these therapies, provided proactive monitoring continues. Integrating WGS into AMR surveillance in resource-limited settings can close knowledge gaps, strengthen outbreak detection, and support One Health strategies for regional and global AMR control. Looking ahead, comparative ARG modelling across Ethiopia, Kenya, and Tanzania could provide predictive insights, enabling early warning systems and targeted interventions to suppress AMR spread.

#### Study Strengths

This study utilized available whole genome sequencing (WGS) datasets compiled from multiple independent sources, enabling access to large data sets. This dataset enabled wide-ranging analysis of *C. jejuni* in different settings and geographical regions, specifically Ethiopia, Tanzania and Kenya. This work was conducted as part of bioinformatic training exercise for Masters’ students, demonstrating the effective application of genomic tools for education and research purposes.

#### Limitations

This study has several limitations. The sampling coverage and number of isolates from Tanzania, Kenya, and Ethiopia were limited, which may not fully represent the regional distribution of antimicrobial resistance. The analysis focused solely on *C. jejuni*, and including additional bacterial species could have provided a broader understanding of resistance dissemination. Moreover, the study relied on genomic data without complementary phenotypic susceptibility testing or functional validation of the detected genes, which may limit interpretation of their actual resistance expression and transfer potential. Finally, incomplete metadata, such as antibiotic usage history and environmental parameters, restricted the ability to link resistance patterns to specific drivers.

## Data Availability

All data produced in the present work are contained in the manuscript

## Author Contributions

JJM, VEL, MAY, YDS, EFM and BL: conceptualization, data curation, methodology. JJM, VEL, MAY, YDS and EFM analyzed and interpreted the data. BL validation and review. JJM, VEL, MAY, YDS, EFM, BL, EK, JM, CK, FR, CS, VT, EM, AM, EB, EM wrote the manuscript, and submitted it for publication. The authors are driven by a commitment to advancing the integration of genomic surveillance into One health approach

## Acknowledgements

The authors grateful acknowledge all research teams and study participants involved in generating the Whole Genome Sequences and submitting them to public repositories which made this analysis possible.

## Conflict of Interest

The authors declare there is no conflict of interest regarding the conduct of this study.

## Ethical Clearance

No ethical clearance was required, as the data were downloaded from the public database.

## Funding

No funds were allocated for the analysis or preparation of this manuscript.

## Notes

### Competing Interest Statement

The authors have declared no competing interest.

### Funding Statement

This study did not receive any funding

### Author Declarations

This study utilized publicly available whole genome sequence (WGS) data of Campylobacter jejuni obtained from the NCBI database, including isolates originating from Homo sapiens, Gallus gallus, Ovis aries, Bos indicus and milk from dairy cattle. No new samples were collected and no direct interaction with human participants occurred.

